# White matter markers of chronic pain and trauma in UK Biobank

**DOI:** 10.64898/2026.05.19.26353622

**Authors:** Tong En Lim, Sylvia M. Gustin, Yann Quidé

## Abstract

**Background:** Lifetime exposure to trauma is associated with chronic pain. Separate studies of chronic pain and trauma report overlapping alterations in white matter microstructure, yet their distinct and cumulative effects remain unclear.

**Methods:** White matter microstructure (fractional anisotropy [FA] and mean diffusivity [MD]) from the UK Biobank (N = 21,995) were analysed using linear mixed-effects models. First, group effects (chronic pain versus control) on white matter integrity within this cohort were established. To investigate distinct and cumulative impacts of trauma exposure at different developmental stages, main and interactive effects of group and trauma severity on FA and MD were examined in separate groups exposed to childhood maltreatment only, adulthood trauma only, and both. Sex-stratified analyses were conducted.

**Results:** Chronic pain was associated with widespread alterations and was spatially refined to brainstem tracts and cingulum when accounting for maltreatment/trauma severity. Accounting for chronic pain, cumulative trauma severity was associated with alterations in brainstem, frontal and parietal tracts, whereas adulthood trauma showed comparable but attenuated patterns. Childhood maltreatment severity was associated with localised FA and MD reductions in brainstem tracts, sagittal stratum and superior longitudinal fasciculus. These effects were more pronounced in females than males. A chronic pain-by-maltreatment/trauma severity interaction was observed for FA in the superior cerebellar peduncle in females exposed to childhood maltreatment only.

**Conclusions:** Distinct and interactive effects of chronic pain and maltreatment/trauma severity on white matter microstructure were evident. The findings suggest that trauma-informed care should be tailored by timing of exposure and sex in this population.

## 1. Introduction

Chronic pain is a major health burden, impacting more than 20% of the global population (1,2). Individuals with chronic pain report approximately twice as much lifetime trauma exposure as the general population (3). When exposed to traumatic experiences, this population often reports more severe pain, functional disability and greater psychological distress (3). Chronic pain and trauma exposure are associated with overlapping alterations in white matter integrity (4,5), but the unique and cumulative effects of chronic pain and trauma exposure remain poorly understood (6).

Alterations in white matter microstructure, including lower fractional anisotropy (FA) and greater mean diffusivity (MD), in tracts including the uncinate fasciculus, corpus callosum and the cingulum bundle, has been reported in individuals with chronic pain (7–9). Importantly, comparable patterns of lower FA and higher MD in these same tracts are also commonly reported in separate studies of individuals exposed to traumatic experiences (10). Despite their co-occurrence and shared brain alterations, very few studies aimed to disentangle the specific and cumulative impacts of chronic pain and trauma exposure on the integrity of white matter (11). More severe post-traumatic stress symptoms have been associated with reduced FA in the uncinate fasciculus in chronic pain-free controls, but not in individuals experiencing chronic pain, possibly indicating different coping mechanisms in pain-free individuals (5). However, the small sample size of this study (N=56, including 36 people with various chronic pain conditions and 20 pain-free controls) may have undermined the discovery of more subtle effects on white matter integrity.

The developmental timing of trauma exposure can differentially impact white matter integrity (12,13). In adults with post-traumatic stress disorder (PTSD), childhood trauma was associated with lower FA in the corpus callosum, whereas trauma exposure in adulthood was associated with both lower and greater FA in the anterior and posterior cingulum, the superior longitudinal fasciculus and frontal regions (10). Despite evidence of overlapping alterations associated with trauma exposure and chronic pain, the timing of trauma exposure on white matter integrity in adults who experience chronic pain remains poorly understood. Additionally, widely established sex differences in white matter microstructure, as well as in the prevalence of chronic pain and trauma exposure, highlight the importance of considering sex differences in the effects of chronic pain and trauma exposure (14).

Leveraging the large dataset from the UK Biobank (UKB), we first characterised the effects of chronic pain on white matter microstructure to clarify inconsistent findings from previous smaller studies, without accounting for trauma exposure. This was followed by sex-stratified analyses to determine sex differences in chronic pain-associated white matter alterations. Next, we examined how the severity of exposure to childhood maltreatment, trauma in adulthood, and their cumulative effects can modulate white matter microstructure differences between adults experiencing chronic pain and pain-free controls. Sex differences in relation to maltreatment/trauma severity were also examined. We hypothesised the severity of trauma exposure to be associated with compromised white matter microstructure integrity (i.e., lower FA and greater MD) in the uncinate fasciculus, cingulum bundle, corpus callosum and superior longitudinal fasciculus, in adults with chronic pain. Differential impacts of trauma depending on the timing of exposure were expected, with cumulative exposure to trauma/maltreatment to be associated with more pronounced effects on white matter microstructure compared to exposure limited to childhood or adulthood alone.

## 2. Methods and materials

### 2.1 Participants

This study conducted secondary analyses using data from the UKB (release v19; application ID 363586). The UKB recruited over 500,000 adults (aged 40-69) and collected extensive phenotypic and genotypic information. The exclusion criteria were: (1) did not attend brain imaging scan at Instance 2, (2) incomplete/missing responses to relevant questionnaires and (3) have medical diagnoses of neoplasm, diseases of the nervous system, obesity (to reduce confounding effects relating to extreme body mass), diabetes and/or narrow-band traumatic brain injury (International Statistical Classification of Diseases and Related Health Problems – version 10 [ICD-10] C00-D48, G00-G99, E65-E68, E10-E14, S00-S09). These ICD-10 diagnoses were excluded to isolate pain-specific supraspinal mechanisms and reduce potential confounding effects arising from brain injury and/or dysfunction.

### 2.2 Demographic information

Demographic information including age, sex and ethnicity was collected. The Townsend deprivation index assessed socioeconomic deprivation (accounting for unemployment, overcrowding, non-car ownership and non-home ownership) (15); higher score represents higher levels of socioeconomic deprivation. Records of ICD-10 mental and behavioural disorders diagnoses (F00-F99) (field-ID 41270) were obtained. Given the high rate of psychiatric comorbidities among participants who experience chronic pain, individuals with diagnosis of mental and behavioural disorders were not excluded; instead, their presence was accounted for in focal analyses.

### 2.3 Chronic pain

Pain experienced in the last month (field-ID 6159) at seven specific body sites (head, face, neck/shoulder, back, stomach/abdomen, hip, knee) or all over the body (multisite) were reported. Participant who responded “none of the above” were included in the control (pain-free) group. Participants reporting pain (in the last month) at any site(s) were then asked whether the pain had persisted for more than 3 months (category-ID 100048). Participants indicating positive responses were classified as having chronic pain, consistent with previous studies (16); those indicating pain lasting for less than 3 months (i.e., experiencing acute pain) were excluded from further analyses.

### 2.4 Childhood maltreatment, trauma exposure in adulthood and cumulative trauma

The Childhood Trauma Screener (CTS-5) was administered to measure severity of childhood maltreatment (17,18). The CTS-5 (field-ID 20487-91) consists of five items describing physical, sexual and emotional abuse, and physical and emotional neglect (for details, see Supplementary Table S1). Participants responded using a 5-point Likert scale from 0 (“Never true”) to 4 (“Very often true”). Two positively worded questions were reverse coded. The items were summed to compute an overall childhood maltreatment severity score (range=0-20; higher score represents higher severity).

An equivalent screener (field-ID 20521-25), adapted from a UK national crime survey for being a victim of crime and adult domestic violence, was used to measure severity of trauma exposure in adulthood (after age 16) (17,19). The screener consists of five items describing relationship and financial insecurity, and physical, psychological and sexual intimate partner violence, measured using a 5-point Likert scale from 0 (“Never true”) to 4 (“Very often true”) (for details, see Supplementary Table S1). Two positively worded questions were reverse coded. Items reflecting the severity of interpersonal trauma exposure (excluding the item reflecting financial insecurity) were summed to compute an overall adulthood trauma severity score (range=0-16; higher score represents higher severity). Both childhood maltreatment and adulthood trauma severity scores were converted to z-scores and summed up to compute an overall score reflecting the cumulative severity of childhood maltreatment and trauma exposure in adulthood (i.e., cumulative trauma).

To examine the distinct and cumulative impacts of maltreatment/trauma severity across developmental periods, childhood maltreatment, adulthood and cumulative trauma were modelled separately. For analyses related to childhood maltreatment, participants who reported no trauma exposure in adulthood (adulthood trauma score=0) were included, and the CTS-5 total score (CTS≥0) was used as an index of childhood maltreatment severity. For analyses related to adulthood trauma, participants who reported no childhood maltreatment exposure (CTS-5=0) were included, and the adulthood trauma screener total score (adulthood trauma score≥0) was used as an index of adulthood trauma severity. For analyses related to cumulative trauma, participants who reported exposure to childhood maltreatment alone (CTS-5>0, adulthood trauma score=0) or trauma in adulthood alone (CTS-5=0, adulthood trauma score>0) were excluded, and standardised cumulative trauma severity score was used.

### 2.5 Imaging acquisition and processing

Tabulated diffusion tensor imaging (DTI) data at first imaging visit were accessed. DTI and T1-weighted structural scans were acquired using 3T Siemens Skyra scanners equipped with 32-channel head coils across four scanning sites (for imaging parameters, see Supplementary Table S2) (20,21). All data underwent minimal pre-processing with Tract-Based Spatial Statistics (22) and quality control using protocols documented in Miller et al. (2016). Indices of white matter microstructure including fractional anisotropy (FA) and mean diffusivity (MD) defined using the Johns Hopkins University tract atlas (23) were extracted.

### 2.6 Statistical analyses

First, linear mixed-effects models were applied to the largest sample of adults with chronic pain possible (irrespective of trauma-related questionnaire availability) to assess the overall effects of chronic pain on white matter microstructure using “*nlme*” package (v3.1-168) (24) in R (v4.4.0) (25). Sex-stratified analyses were performed to examine sex differences; regression coefficients for the group differences were compared between male and female models using Z-tests. Exploratory analyses in the larger sample were conducted to characterise white matter alterations associated with chronic pain across different body sites. Second, linear mixed-effects models were applied to examine main effects and interactions between chronic pain and trauma/maltreatment severity (the product of group by mean-centred trauma/maltreatment severity) separately for participants exposed to childhood maltreatment only, adulthood trauma only and both. Sex-stratified analyses were also performed; regression coefficients for the main and interaction effects were compared between male and female models using Z-tests. In all analyses, subject ID and imaging sites were modelled as random effects and covariates included age, squared mean-centred age (to account for non-linear effects of age), sex, Townsend deprivation index and presence of mental health diagnoses. Outcomes with extreme residuals (absolute skewness > 2 or kurtosis > 10) were winsorised at the 5th and 95th percentiles and refitted to reduce the influence of potential outliers. False discovery rate (FDR) correction using Benjamini-Hochberg method (26) was applied to account for the number of regions-of-interest (ROIs) studied (48 ROI tracts). Statistical significance was set at *FDR-corrected p*-value (*pFDR)<0.05*. Effect sizes were reported as Cohen’s d for group-level chronic pain comparisons and as standardised beta for associations with maltreatment/trauma severity.

## 3. Results

### 3.1 Participant characteristics

Demographic characteristics of the larger UKB chronic pain sample (N=36,955) are presented in Supplementary Table S3, and demographic characteristics for subset of participants who completed trauma-related questionnaires (N=21,995) are summarised in Table 1. The control group included 13,248 participants (60.2%; 52.6% female) while the chronic pain group included 8,747 participants (39.8%; 58.6% female). Multisite pain (50.5%) was most reported, followed by knee (16.7%) and back (10.4%) pain. Within the control group, 18.9% (46.8% female) reported exposure to childhood maltreatment only, 19.3% (57.5% female) to adulthood trauma only, and 37.6% (54.0% female) to both. Within the chronic pain group, 18.4% (48.6% female) reported exposure to childhood maltreatment only, 17.3% (65.6% female) to adulthood trauma only and 44.5% (61.1% female) to both. Relative to controls, the chronic pain group was significantly younger, reported being exposed to more severe trauma/maltreatment (childhood, adulthood and cumulative), included more females and a higher proportion of participants with diagnoses of psychiatric comorbidities.

**Table 1:**
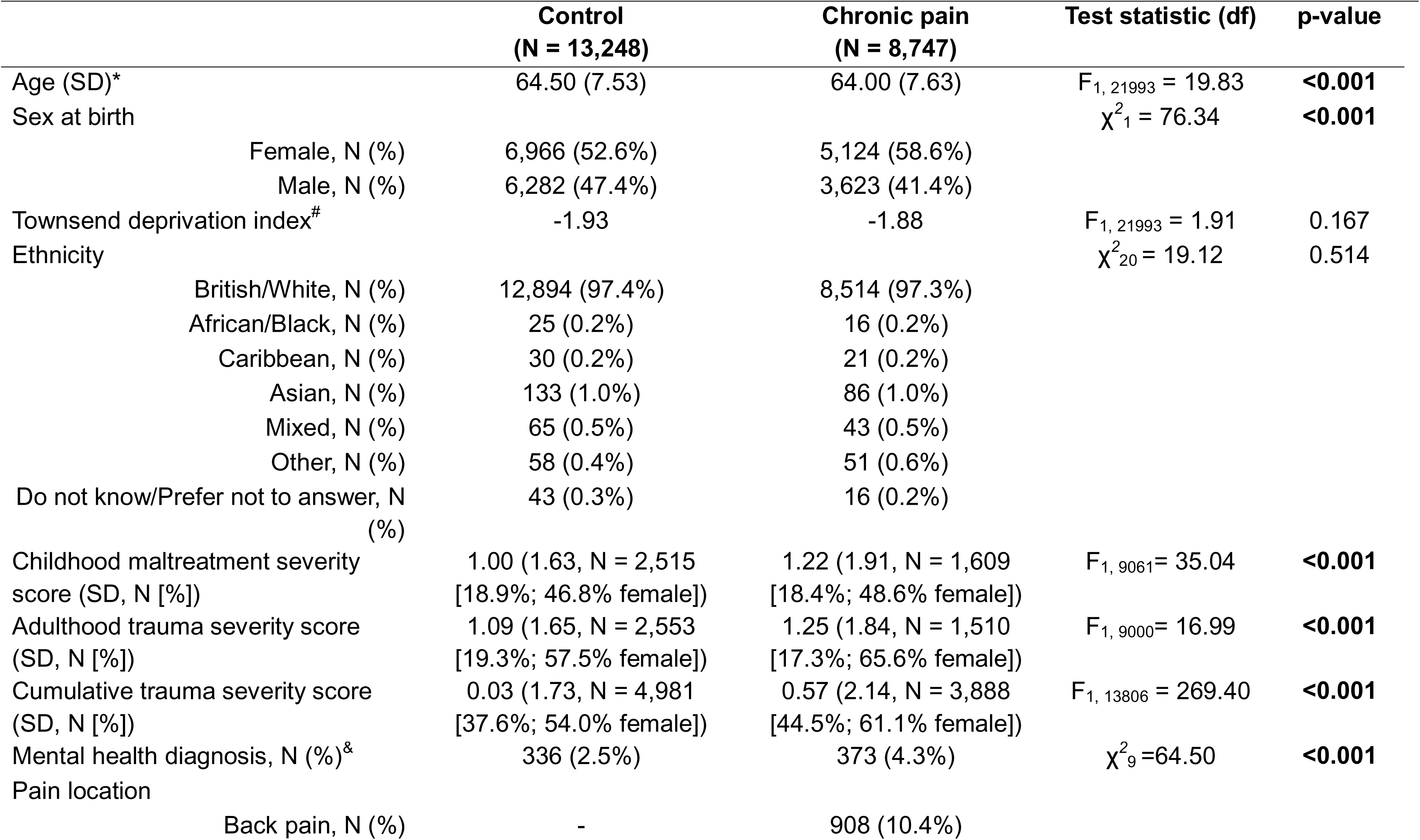
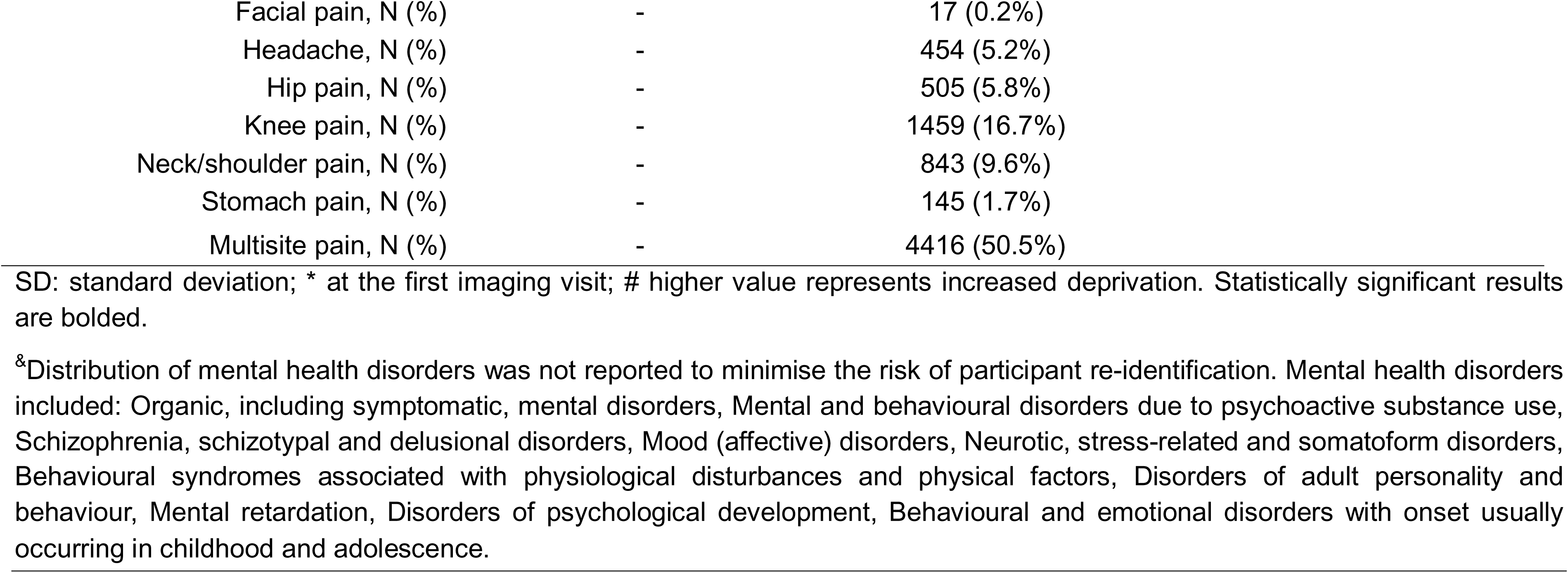
Demographic information.

| Table 1: Demographic information |  |  |  |  |
| --- | --- | --- | --- | --- |
|  | Control<br>(N = 13,248) | Chronic pain<br>(N = 8,747) | Test statistic (df) | p-value |
| Age (SD)* | 64.50 (7.53) | 64.00 (7.63) | $F_{1, 21993} = 19.83$ | **<0.001** |
| Sex at birth | | | $\chi^2_{1} = 76.34$ | **<0.001** |
| Female, N (%) | 6,966 (52.6%) | 5,124 (58.6%) |  |  |
| Male, N (%) | 6,282 (47.4%) | 3,623 (41.4%) |  |  |
| Townsend deprivation index<sup>#</sup> | -1.93 | -1.88 | $F_{1, 21993} = 1.91$ | 0.167 |
| Ethnicity | | | $\chi^2_{20} = 19.12$ | 0.514 |
| British/White, N (%) | 12,894 (97.4%) | 8,514 (97.3%) |  |  |
| African/Black, N (%) | 25 (0.2%) | 16 (0.2%) |  |  |
| Caribbean, N (%) | 30 (0.2%) | 21 (0.2%) |  |  |
| Asian, N (%) | 133 (1.0%) | 86 (1.0%) |  |  |
| Mixed, N (%) | 65 (0.5%) | 43 (0.5%) |  |  |
| Other, N (%) | 58 (0.4%) | 51 (0.6%) |  |  |
| Do not know/Prefer not to answer, N (%) | 43 (0.3%) | 16 (0.2%) |  |  |
| Childhood maltreatment severity score (SD, N [%]) | 1.00 (1.63, N = 2,515 [18.9%; 46.8% female]) | 1.22 (1.91, N = 1,609 [18.4%; 48.6% female]) | $F_{1, 9061} = 35.04$ | **<0.001** |
| Adulthood trauma severity score (SD, N [%]) | 1.09 (1.65, N = 2,553 [19.3%; 57.5% female]) | 1.25 (1.84, N = 1,510 [17.3%; 65.6% female]) | $F_{1, 9000} = 16.99$ | **<0.001** |
| Cumulative trauma severity score (SD, N [%]) | 0.03 (1.73, N = 4,981 [37.6%; 54.0% female]) | 0.57 (2.14, N = 3,888 [44.5%; 61.1% female]) | $F_{1, 13806} = 269.40$ | **<0.001** |
| Mental health diagnosis, N (%)<sup>&</sup> | 336 (2.5%) | 373 (4.3%) | $\chi^2_{9} = 64.50$ | **<0.001** |
| Pain location |  |  |  |  |
| Back pain, N (%) | - | 908 (10.4%) |  |  |

|  |  |  |
| --- | --- | --- |
| Facial pain, N (%) | - | 17 (0.2%) |
| Headache, N (%) | - | 454 (5.2%) |
| Hip pain, N (%) | - | 505 (5.8%) |
| Knee pain, N (%) | - | 1459 (16.7%) |
| Neck/shoulder pain, N (%) | - | 843 (9.6%) |
| Stomach pain, N (%) | - | 145 (1.7%) |
| Multisite pain, N (%) | - | 4416 (50.5%) |

SD: standard deviation; \* at the first imaging visit; # higher value represents increased deprivation. Statistically significant results are bolded.
<sup>&</sup>Distribution of mental health disorders was not reported to minimise the risk of participant re-identification. Mental health disorders included: Organic, including symptomatic, mental disorders, Mental and behavioural disorders due to psychoactive substance use, Schizophrenia, schizotypal and delusional disorders, Mood (affective) disorders, Neurotic, stress-related and somatoform disorders, Behavioural syndromes associated with physiological disturbances and physical factors, Disorders of adult personality and behaviour, Mental retardation, Disorders of psychological development, Behavioural and emotional disorders with onset usually occurring in childhood and adolescence.

### 3.2 Chronic pain-related white matter alterations

Statistical details for the analyses investigating the effects of chronic pain on white matter metrics using the larger sample, irrespective of trauma-related data availability, are reported in Supplementary Table S4. Relative to controls, chronic pain was associated with widespread FA and MD alterations across major white matter tracts, including the corpus callosum, internal capsule, corona radiata, cerebellar pathways, brainstem, thalamic radiations, and fronto-occipital and cingulum tracts (see Figure 1A). Sex-stratified analyses revealed that females show more widespread chronic pain-associated alterations, including lower FA and MD in the frontal, parietal, cingulum and brainstem tracts and corpus callosum, and greater MD in the right superior frontal-occipital fasciculus; males show lower FA and MD in the brainstem tracts, sagittal stratum and corpus callosum (see Figure 1B). However, these differences between sexes were not statistically significant (see Supplementary Table S5). Exploratory analyses examining white matter microstructure differences by pain location relative to controls are presented in Supplementary Table S6.

**Figure 1.**
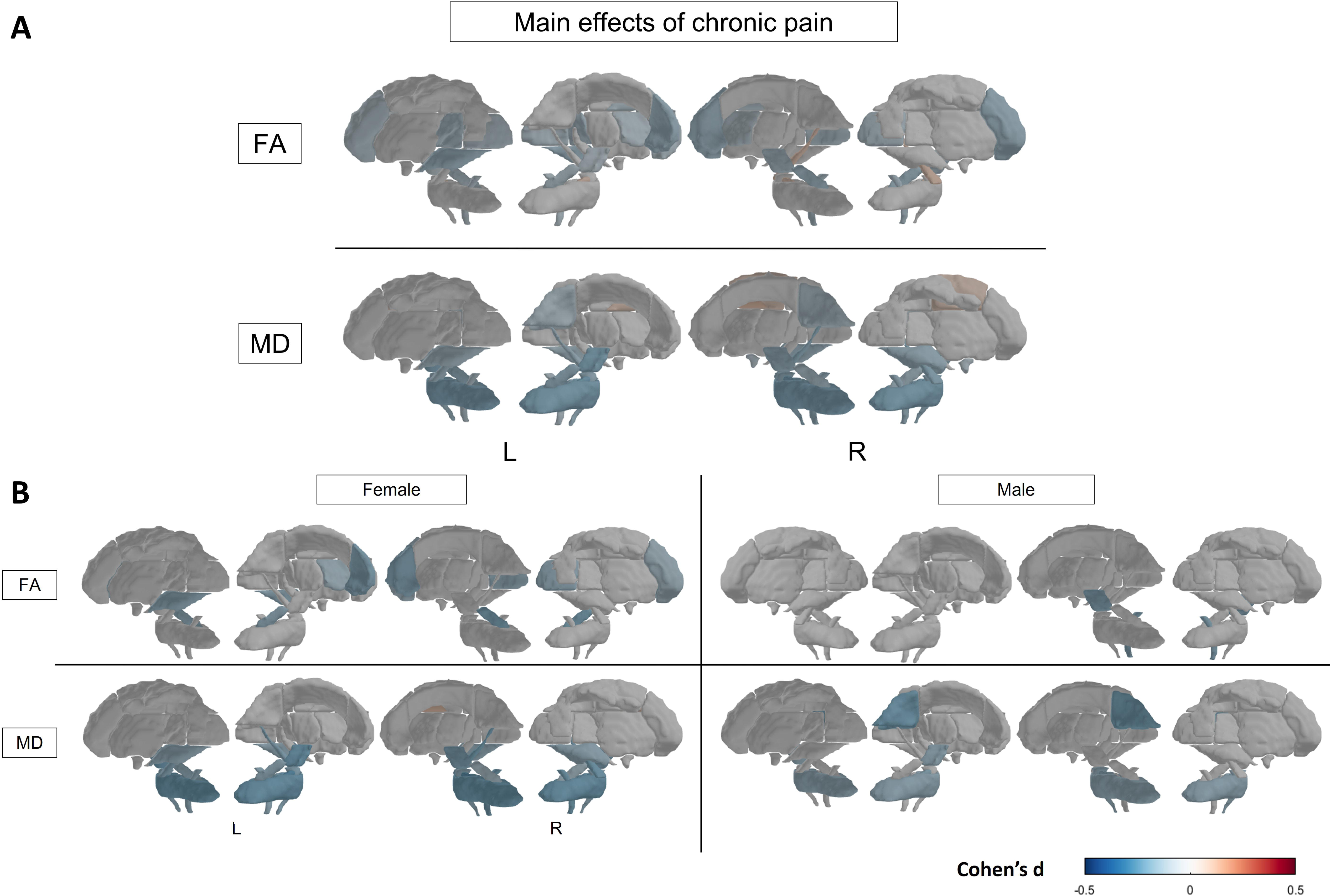
Significant effects of chronic pain on white matter microstructure metrics in the larger UKB chronic pain sample. (A) Significant differences in fractional anisotropy (FA) and mean diffusivity (MD) associated with chronic pain. (B) Significant differences in FA and MD in females (left panel) and males (right panel). Effects sizes are presented Cohen’s d; positive effect size (red) represents increase whereas negative effect size (blue) represents decrease.

### 3.3 Childhood maltreatment

Statistical details for the analyses investigating the effects of group, severity of childhood maltreatment and their interactions on white matter metrics are presented in Supplementary Tables S7 and S8. When accounting for the severity of childhood maltreatment, chronic pain was associated with lower FA in the right cerebral peduncle compared to controls (see Figure 2A). Severity of childhood maltreatment was associated with lower FA in the sagittal stratum, superior and inferior cerebellar peduncle, superior longitudinal fasciculus, cerebral peduncle, medial lemniscus, as well as lower MD in the corticospinal tract, when accounting for group status. There were no significant differences between sexes in the effects of chronic pain and childhood maltreatment severity (see Supplementary Table S9 and S10). The group-by-maltreatment severity interaction was significantly associated with FA in the right superior cerebellar peduncle in females only (see Figure 2B). Specifically, more severe childhood maltreatment was associated with increased FA in the right superior cerebellar peduncle in females with chronic pain relative to controls, but not in males.

**Figure 2.**
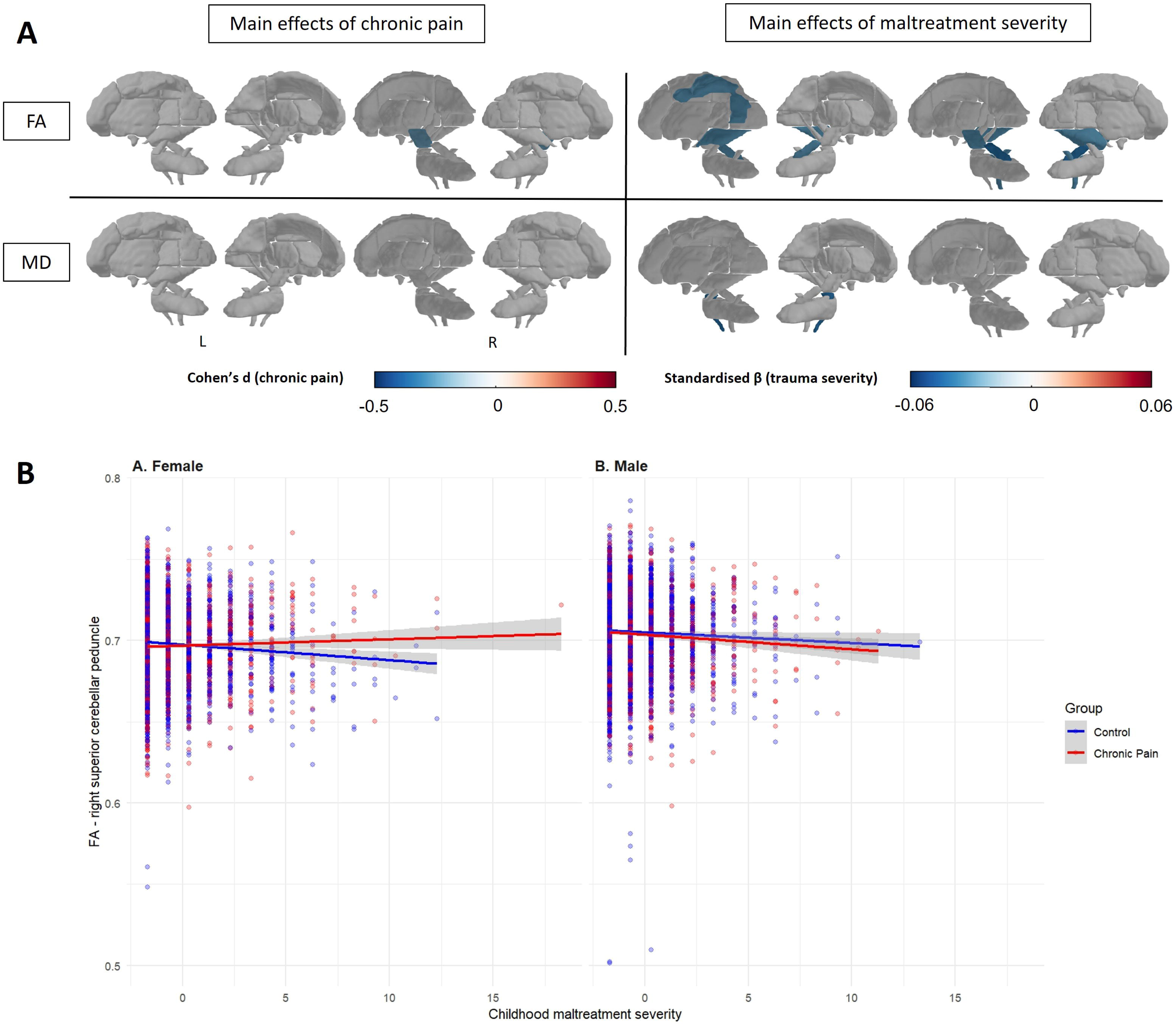
Significant effects of chronic pain and severity of childhood maltreatment exposure on white matter microstructure metrics. (A) Significant differences in fractional anisotropy (FA) and mean diffusivity (MD) associated with chronic pain (left panel) and severity of childhood maltreatment exposure (right panel). (B) Group-by-childhood maltreatment severity interactions associated with FA of the right superior cerebellar peduncle in females (left panel) and males (right panel) Effects sizes are presented in standardised beta (trauma severity) and Cohen’s d (chronic pain); positive effect size (red) represents increase/positive relationship whereas negative effect size (blue) represents decrease/negative relationship.

### 3.4 Trauma exposure in adulthood

Statistical details for the analyses investigating the effects of group, severity of adulthood trauma exposure and their interactions on white matter metrics are presented in Supplementary Tables S11 and S12. Severity of adulthood trauma exposure was significantly associated with widespread lower FA in the corpus callosum, limbic pathways, internal capsule, cerebellar tracts, brainstem and association fibres such as the superior longitudinal and fronto-occipital fasciculus, when accounting for group status (see Figure 3A). Adulthood trauma severity was also significantly associated with greater MD in the anterior corona radiata, cingulum (hippocampus) and fornix cres stria terminalis. Group and group-by-trauma severity interaction were not significantly associated with variations in any of the ROIs studied. Sex-stratified analyses showed that more severe trauma exposure in adulthood was associated with lower FA in the cingulum, parietal and occipital, brainstem tracts and corpus callosum, and greater MD in the fornix cres stria terminalis in females; no alterations were observed in males (see Figure 3B). The differences between sexes, however, were not statistically significant (see Supplementary Table S13 and S14).

**Figure 3.**
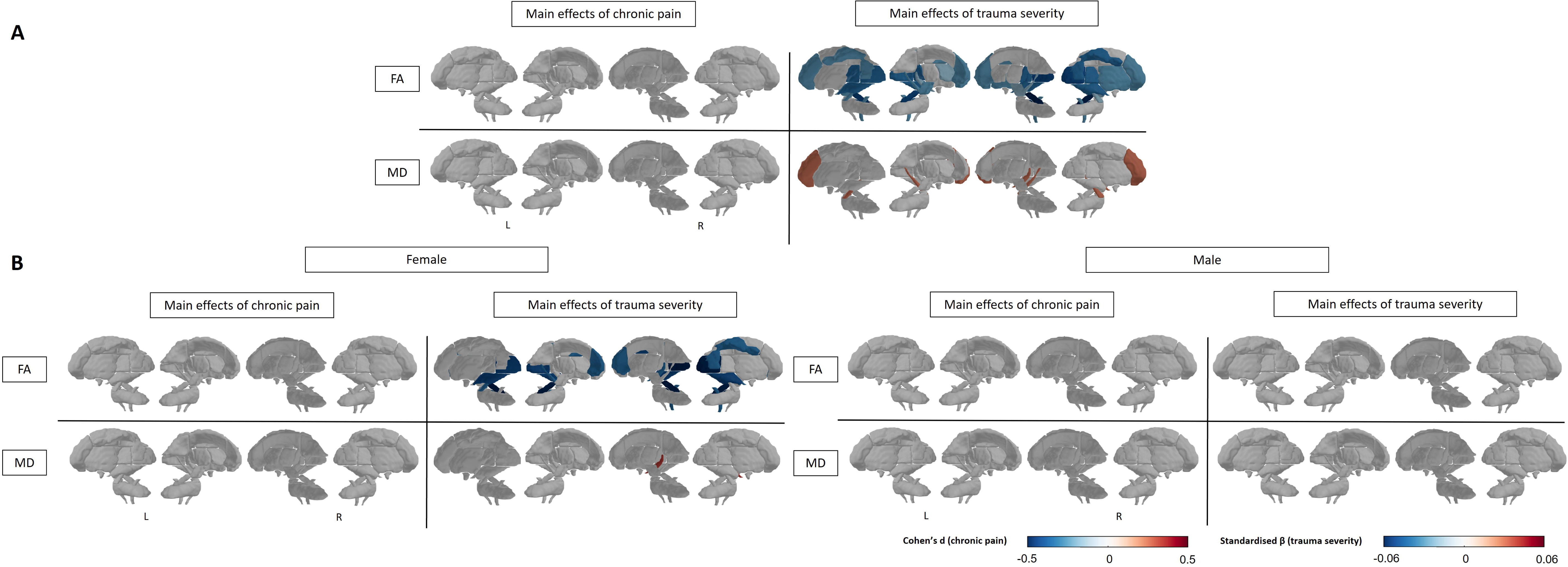
Significant effects of chronic pain and severity of trauma exposure in adulthood on white matter microstructure metrics. Significant differences in fractional anisotropy (FA) and mean diffusivity (MD) associated with chronic pain (left panel) and severity of trauma exposure in adulthood (right panel). (B) Significant chronic pain- and trauma-associated differences in FA and MD in females (left panel) and males (right panel). Effects sizes are presented in standardised beta (trauma severity) and Cohen’s d (chronic pain); positive effect size (red) represents increase/positive relationship whereas negative effect size (blue) represents decrease/negative relationship.

### 3.5 Cumulative trauma

Statistical details for the analyses investigating the effects of group, cumulative trauma severity and their interactions on white matter metrics are presented in Supplementary Tables S15 and S16. When accounting for the severity of cumulative trauma, chronic pain was associated with lower MD across cerebellar, brainstem, and projection tracts, including the middle cerebellar, pontine crossing, cerebral peduncle, cingulum, corticospinal, and cerebellar peduncle pathways, and greater MD in the bilateral superior fronto-occipital fasciculus (see Figure 4A). Severity of cumulative trauma was associated with lower FA across widespread white matter tracts, including interhemispheric, limbic, projection, cerebellar and association pathways such as the corpus callosum, fornix, internal capsule, cingulum, corona radiata, thalamic radiations, brainstem tracts and major association fibres, when accounting for group status. In addition, severity of cumulative trauma was associated with greater MD in the genu corpus callosum, corona radiata, and posterior thalamic radiations and superior longitudinal fasciculus, as well as with lower MD in the pontine crossing and corticospinal tracts. Group-by-trauma severity interaction was not significantly associated with variation in any of the ROIs studied. Sex-stratified analyses revealed that, in females, cumulative trauma severity was associated with widespread lower FA and lower or greater MD, whereas in males, cumulative trauma severity was associated with lower FA in the cerebellar peduncle and parts of the frontal and parietal tracts. Chronic pain was associated with lower FA in the brainstem tracts in females, and no chronic pain-related alterations were observed in males (see Figure 4B). The differences between sexes were not statistically significant (see Supplementary Table S17 and S18).

**Figure 4.**
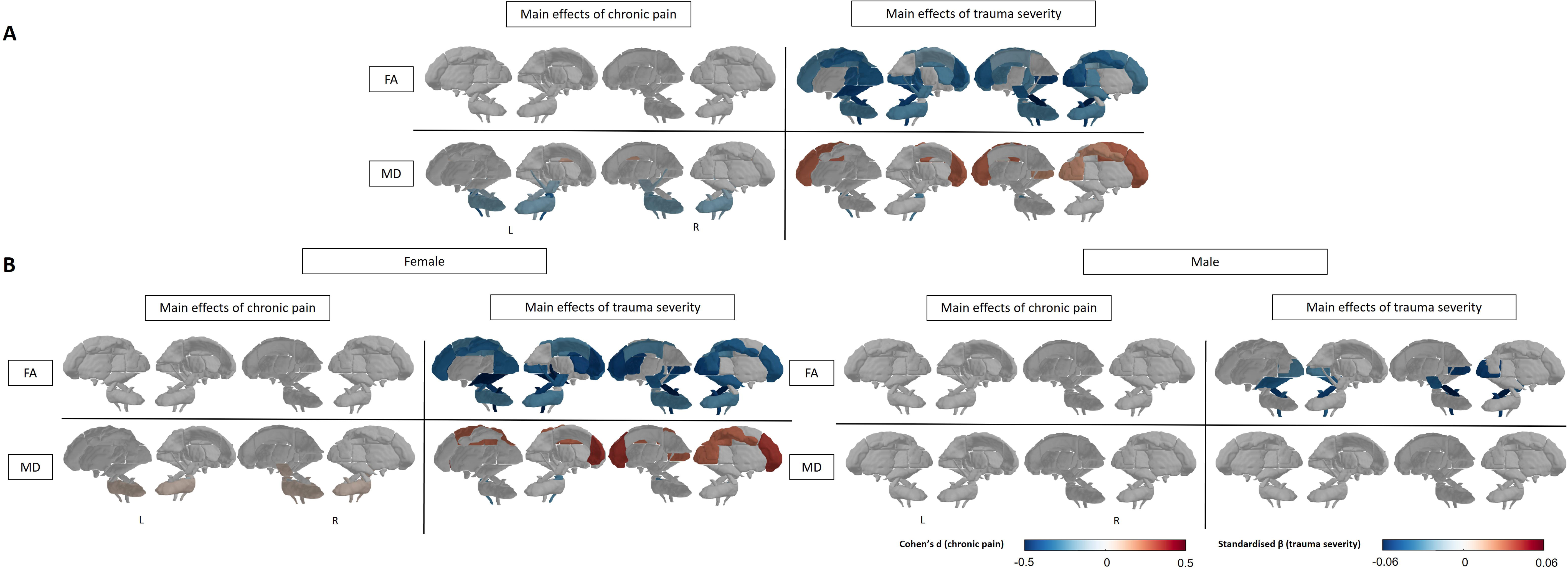
Significant effects of chronic pain and cumulative severity of childhood maltreatment and trauma exposure in adulthood on white matter microstructure metrics. Significant differences in fractional anisotropy (FA) and mean diffusivity (MD) associated with chronic pain (left panel) and cumulative severity of childhood maltreatment and trauma exposure in adulthood (right panel). (B) Significant chronic pain- and trauma-associated differences in FA and MD in females (left panel) and males (right panel). Effects sizes are presented in standardised beta (trauma severity) and Cohen’s d (chronic pain); positive effect size (red) represents increase/positive relationship whereas negative effect size (blue) represents decrease/negative relationship.

## 4. Discussion

The present study established white matter microstructural markers of chronic pain in the UKB, and how the severity of childhood maltreatment, adulthood trauma or their cumulative impacts contribute to these effects. Chronic pain overall was associated with widespread FA and MD alterations compared to controls; when accounting for maltreatment/trauma severity, chronic pain-related alterations were localised in the brainstem and cingulum. Severity of adulthood and cumulative trauma were associated with widespread white matter alterations, whereas severity of childhood maltreatment, when no later trauma exposure was reported, was associated with FA and MD reductions localised to the brainstem, sagittal stratum and superior longitudinal fasciculus. Relative to males, females show more pronounced chronic pain- and trauma-related white matter alterations. Chronic pain-by-maltreatment/trauma severity interactions were associated with FA in the right cerebellar peduncle in females, but not males, exposed to childhood maltreatment only. These findings suggest that effects of chronic pain and maltreatment/trauma severity were different among sexes and timing of trauma exposure, highlighting the importance of incorporating these factors into interventions for this population.

Relative to controls, individuals with chronic pain showed widespread FA and MD alterations across frontal, parietal, occipital regions as well as the corpus callosum, cingulum, brainstem and cerebellar tracts. There were no significant sex differences in any indices of white matter microstructure; however, female exhibited alterations overlapping with those observed in males, including the brainstem and corpus callosum, but with larger effect size and broader spatial involvement, extending into frontal pathways and sagittal stratum. Current evidence on white matter microstructural alterations in chronic pain remains largely inconclusive: increased, decreased or unchanged FA and MD have been reported in the cingulum bundle, corpus callosum, motor pathways, frontal and parietal tracts, and brainstem across chronic pain conditions (i.e., chronic musculoskeletal pain, fibromyalgia) (7,27). These conflicting results may be partly due to small sample sizes (minimum N=7, maximum N=74), sex and/or pain location/condition, as suggested by our exploratory analyses (5,27). Variability in chronic pain duration and intensity, as well as the use of medications, may also contribute to these discrepancies (28). By leveraging a large sample and examining sex and pain location differences, the present study provides a more comprehensive characterisation of white matter microstructural alterations associated with chronic pain, that could be targeted for interventions such as transcranial magnetic or direct current stimulations.

When accounting for trauma severity, chronic pain-associated alterations were spatially refined, primarily in the brainstem, cingulum, and superior fronto-occipital fasciculus which are critical for integrating sensory, cognitive and affective aspects of chronic pain including memory, executive control, motor and multisensory processing, pain modulation, and attention (29–32). These alterations may reflect axon/myelin loss, cellular swelling and/or neuroinflammation in these tracts (7,33), and weaken interregional communication (34). These results suggest that some of the variance in white matter microstructure alterations previously attributed to chronic pain overlaps with those related to trauma exposure. Therefore, accounting for trauma allows finer identification of white matter changes associated with chronic pain that may be targeted for interventions.

When accounting for chronic pain, trauma/maltreatment exposure at different timing have different impacts on white mater microstructure, partially consistent with previous studies that did not account for preceding or subsequent trauma exposure across other developmental periods (10,35–38). First, severity of childhood maltreatment was associated with localised FA and MD reductions in the brainstem tracts, sagittal stratum and superior longitudinal fasciculus. On the other hand, severity of adulthood and cumulative trauma were associated with widespread FA reductions and either greater or lower MD in the frontal, parietal and brainstem tracts. These results suggest that childhood maltreatment, in the absence of later trauma exposure, may impact the integrity of tracts supporting critical functions during neurodevelopment, including visual processing, nociception and cognitive control, whereas adulthood and cumulative trauma may have a more widespread impact in tracts critical for emotion processing, cognition control and sensorimotor function. Sex differences in the effects of maltreatment/trauma severity across developmental periods were observed. Reduced white matter integrity was primarily observed in tracts associated with sensory and integrative functions in males, whereas females showed more widespread involvement extending to frontal, cingulum, and interhemispheric tracts implicated in higher-order cognitive and emotional processing. Importantly, no significant childhood maltreatment-associated alterations were observed in either sex, suggesting greater overlap with chronic pain-related effects. This pattern may reflect sex differences in stress responsivity, potentially influenced by gonadal hormones and engagement of affective and regulatory networks (39–41).

A dose-response relationship was evident between trauma exposure and white matter integrity, while accounting for chronic pain. Relative to controls, severity of cumulative trauma exposure, was associated with more pronounced alterations when compared to effects of childhood maltreatment or adulthood trauma alone. This finding is consistent with evidence for elevated allostatic loads, or the cumulative burden of chronic stress on brain integrity (42). The accumulation of exposure to childhood maltreatment and later life traumatic events may lead to heightened allostatic load that can damage white matter integrity, as previously shown in the UKB cohort (43). However, this interpretation remains speculative and will need to be formally tested in future studies. Interestingly, recency effects of trauma exposure, with severity of adulthood trauma alone having more pronounced effects than childhood maltreatment alone, were observed. This potentially reflects specific developmental trajectories of white matter integrity which reaches peak maturation later in life relative to grey matter maturation, between 20 and 42 years of age (44,45). White matter microstructural plasticity may therefore occur and normalise, or at least minimise, the long-term effects of childhood maltreatment on white matter integrity, in the absence of subsequent trauma exposure, compared to those first exposed to trauma in adulthood (46). Finally, chronic pain-by-maltreatment/trauma severity interactions, while not significantly associated with white matter variations in the overall trauma-related samples, were associated with FA in the right superior cerebellar peduncle, a tract critical for motor functions, in females exposed to childhood maltreatment alone. The observed interaction suggests that chronic pain may also act as a stressor that interacts with early-life maltreatment to influence white matter microstructure later in life, potentially reflecting a latent vulnerability in females.

The present study has potential clinical implications. First, the findings suggest that adults exposed to maltreatment/trauma at different developmental periods, while accounting for chronic pain, may benefit from distinct forms of trauma-informed care. Specifically, adults reporting childhood maltreatment without subsequent trauma exposure may benefit from interventions targeting sensory and cognitive control processes, such as physical exercise (47,48) and mindfulness-based (49) approaches. In contrast, adults exposed to adulthood or cumulative trauma may benefit from more integrated interventions targeting broader cognitive-emotional and regulatory processes, such as dialectical behavioural therapy (50,51). Importantly, relative to males, females show more pronounced effects of chronic pain and maltreatment/trauma, mirroring patterns typically associated with greater trauma severity and/or chronic pain, highlighting the need for earlier and/or more integrated interventions in females.

This study has several limitations. First, the study is restricted by the white matter microstructure metrics availability in the UKB dataset. The UKB did not include measures of axial diffusivity, a marker of axonal damage, and radial diffusivity, a marker of myelin damage, for whom FA is a function of (9). These metrics can inform whether observed alterations are driven by axonal damage or/and demyelination. Second, trauma exposure was assessed retrospectively, which may be subject to recall bias, although previous studies suggested that self-report indices of trauma exposure demonstrate good reliability, even in samples with severe psychiatric conditions (52,53). Third, this study did not account for the use of pharmacological treatments that may influence brain integrity (54). Finally, the UKB cohort was recruited from the general population, likely reporting less severe indices of psychopathology, including trauma exposure, than medically diagnosed chronic pain patients, as reflected by the limited range and skewed distribution of trauma severity (see Supplementary Figure S1). Future studies of large clinical populations using a wider array of white matter microstructure metrics, such as data pooled together from the Enhancing NeuroImaging Genetics through Meta-Analysis (ENIGMA)-Chronic Pain working group (55), are needed to better understand the differential effects of trauma/maltreatment exposure in this population.

The present study provides evidence for white matter alterations associated with chronic pain, as well as with childhood maltreatment, adulthood and cumulative trauma. Specifically, chronic pain was associated with widespread white matter alterations; whereas adulthood and cumulative trauma severity was associated with alterations in brainstem, frontal and parietal tracts, and childhood maltreatment-related alterations were localised in the brainstem, sagittal stratum and superior longitudinal fasciculus. Importantly, sex and timing differences in distinct and interactive effects between chronic pain and maltreatment/trauma severity were observed. Females showed more pronounced white matter alterations related to both chronic pain and maltreatment/trauma severity across developmental periods. Additionally, chronic pain-by-childhood maltreatment severity interaction was associated with FA of the superior cerebellar peduncle in females, but not males. Together, the finding suggests that interventions for chronic pain may benefit from integrating trauma-informed care, accounting for the timing of trauma exposure and sex, to improve outcomes.

## Acknowledgements

This research has been conducted using the UKB resource (application no. 363586). Tong En Lim was supported by a University of New South Wales (UNSW Sydney) International Postgraduate Scholarship and Edward C. Dunn scholarship administered by Neuroscience Research Australia (NeuRA). Sylvia M. Gustin was supported by Rebecca Cooper Fellowship from the Rebecca L. Cooper Medical Research Foundation.LLWe are grateful to the study participants for their time and participation.

## Data availability

The data from the UKB used in this study are publicly available via their standard data access procedure at https://www.ukbiobank.ac.uk/.

## Authors’ contribution

Tong En Lim contributed conceptualisation, data curation, formal analysis, methodology, visualization, validation, writing of the original draft, review and editing. Sylvia M. Gustin contributed conceptualisation, funding acquisition, methodology, project administration, resources, supervision, and review and editing. Yann Quidé contributed conceptualisation, data curation, formal analysis, methodology, visualisation, supervision, validation, writing of the original draft, review and editing.

## Declarations of interest

All authors report no biomedical financial interests or potential conflicts of interest.

## Notes

### Competing Interest Statement

The authors have declared no competing interest.

